# Preference for Social Motion in Autistic Adults

**DOI:** 10.1101/2024.02.14.24302687

**Authors:** Magdalena Matyjek, Nico Bast, Salvador Soto Faraco

## Abstract

Autism is often linked to attenuated social attention, including a lowered looking preference to biological motion in autistic compared to non-autistic children. This looking preference has been suggested as an autism marker in childhood. However, few studies have investigated whether this bias persists into adulthood. Furthermore, the underlying cognitive mechanism of this group difference is largely unknown. Pupillary responses have been established as an index of salience processing and are thus a promising measurement of the cognitive bases of looking preference. The present study examined differences in looking preference and pupillary responses to social versus geometric motion between autistic and non-autistic adults (N=66). In terms of preference, autistic adults demonstrated a reduced spontaneous looking toward social stimuli compared to the non-autistic group. Whereas the former displayed no clear preference for either motion type, the latter showed a strong preference for social motion. In terms of pupillary responses, the autistic group showed faster and larger pupil dilation for social motion compared to the non-autistic group, consistent with heightened cognitive effort. These results suggest persistent differences in social attention across the developmental lifespan in autism.

**LAY SUMMARY:** Autism is often associated with differences in social attention, and how much autistic children look at social motion (moving human faces) over non-social motion (geometric shapes) could be applied to screen and prioritise potentially autistic persons for diagnosis and support services. Our study investigated whether these differences persist into adulthood. We found that adult autistic participants spontaneously looked less at video-clips containing social motion compared to their non-autistic counterparts, and showed faster and larger pupillary responses to social motion, which could be an indication of increased cognitive effort in interpreting social information. These findings suggest that the lowered social motion preference in autism persists across lifespan.

## 1. INTRODUCTION

### 1.1. Background

Autism is a life-long neurodevelopmental condition characterised by social and communication difficulties alongside repetitive and restricted behaviours (American Psychiatric Association, 2013). It is often associated with attenuated social attention (Chevallier et al., 2012; Frazier et al., 2017) and atypical processing of human biological (natural movements produced by humans, e.g., walking, sitting, dancing) and social motion (including higher-order information, e.g., emotional facial expressions). While non-autistic individuals have a bias towards biological motion from early infancy (Farroni et al., 2007), this bias is reduced in autistic individuals across lifespan (Federici et al., 2020; Todorova et al., 2019; Van der Hallen et al., 2019). Because biological motion processing is a crucial component of social perception, this reduction may have cascading effects on social development (Pavlova, 2012) and may underlie social difficulties in autism. Thus, researchers have explored biological /social motion preference as a potential autism marker, especially in children (Pierce et al., 2011). A social motion preference task might be widely applied to screen potentially autistic persons for diagnosis and support services. However, it remains to be investigated whether attenuated social motion preference in autism persists into adulthood. Thus, in this study, we investigated social motion preference in autistic and neurotypical adults.

The preferential looking paradigm with social vs. non-social stimuli presented side-by-side is often used to assess participants’ looking preferences. Studies measuring looking preference with naturalistic stimuli consistently demonstrated reduced preference for social and increased preference for geometric motion in autistic toddlers, children, and adolescents, compared to their non-autistic peers (Franchini et al., 2016, 2017; Hong et al., 2019; Moore et al., 2018; Pierce et al., 2011, 2016; Polzer et al., 2022; Shaffer et al., 2017; Vargas-Cuentas et al., 2017; Wen et al., 2022). Furthermore, biological / social motion preference has been positively correlated with adaptive functioning, and negatively with symptom intensity in autistic toddlers and children (Bacon et al., 2020; Franchini et al., 2016, 2017; Moore et al., 2018; Pierce et al., 2016). Although few studies have investigated looking preference in adults, some showed reduced social motion preference in autism (Del Valle Rubido et al., 2020; Fujioka et al., 2016) and in higher autistic traits (Le et al., 2020).

Despite evidence for the reduced preference for biological and social motion in autism is robust, the understanding of its cognitive and neuronal underpinnings is less well understood (Mason et al., 2021). A potential candidate for atypical visual preferences in autism is altered salience processing (Polzer et al., 2022; Zhao et al., 2022). Pupil responses have been linked to the activity of the locus coeruleus-norepinephrine (LC-NE) system (Joshi et al., 2016; Megemont et al., 2022), as a correlate of physiological arousal (Sara & Bouret, 2012) and sensory reactivity to external stimuli based on their salience (McBurney-Lin et al., 2019). The study by Polzer et al. (2022) showed that while non-autistic children looked relatively more often at social motion stimuli, which elicited an increase in their pupil sizes, autistic children showed the opposite pattern: they looked relatively more often and showed larger pupil sizes to geometric motion. To the best of our knowledge, no study has so far investigated if similar pupil responses related to social motion preference persist in adult individuals diagnosed with autism. Answering this question is important because it would provide insights into the developmental trajectory of social perception in autism and shed light on potential differences in cognitive processing between autistic children and adults.

### 1.2. The current study

We investigate social motion preference in autistic adults, compared to age, gender, and intelligence-matched non-autistic peers without diagnosed neuro-or psychological conditions (henceforth, neurotypicals). For the looking preference (task 1), we adapted the preferential looking paradigm typically used in children (Pierce et al., 2011). Participants watched side-by-side videos with one side showing actors producing facial expressions (social; SOC) and the other side showing dynamic geometric patterns (GEO). Task 2 was designed to measure pupil dilation without eye movements and limiting strategic control, because adults differ from young children in gaze patterns in free viewing (Franchak et al., 2016) and their overt orienting (or lack thereof) cannot be directly interpreted as preference. For instance, adults may have developed compensatory behaviours, and /or mentalise about the study’s hypothesis and experimenters’ expectations. Thus, in task 2, we recorded participants’ pupil sizes while they passively watched SOC and GEO stimuli presented one at a time in the centre of the monitor.

### 1.3. Hypotheses and predictions

We hypothesised that if, similarly to children, autistic and non-autistic adults differ in social attention, then similar group differences in social vs. geometric motion preference and physiological responses would be observed. Specifically, we predicted that:

- **Hypothesis 1:** the autistic (AUT) compared to neurotypical (NT) group would spend relatively more time looking at geometric than social stimuli when presented simultaneously. If so, we planned to test whether NT and AUT exhibit a specific preference for social and geometric motion, respectively.
- **Hypothesis 2:** AUT versus NT have relatively larger pupil sizes when passively watching geometric than social motion. If so, we would further test whether NT show larger pupil sizes to social stimuli and AUT to non-social stimuli.

The hypotheses, sampling plan, and analyses were pre-registered at https://osf.io/sp2gc prior to any data analyses.

## 2. METHOD

### 2.1. Sample size determination

Sample size was targeted at N=35 per group, as dictated by the requirements of concurrent tasks in the data collection session. With the total sample of N=70, a regression model assessing looking preference with one predictor (group), statistical power β=0.8, at an α level=0.05, allows for the detection of a medium effect size of f^2^ =0.11.

### 2.2. Participants

We recruited 35 autistic and 41 non-autistic adult Spanish-speaking volunteers with normal or corrected-to-normal vision and hearing, without intellectual disability (IQ score>80 in the Raven’s Progressive Matrices 2; John & Raven, 2003), motor tics, or seizures. Ten participants were excluded following pre-registered criteria (see details below) and the final sample size was 32 autistic and 34 non-autistic participants.

Autistic (AUT) participants were recruited through invitations shared with autism centres in Barcelona, social media, flyers at public events, and internal communication within the local autism community. All AUT had a confirmed diagnosis of autism spectrum disorder made by professionals in specialised autism centres in Catalonia after 2013, in accordance with DSM-5 criteria (American Psychiatric Association, 2013). Of these, 24 participants had undergone the Autism Diagnostic Observation Schedule (ADOS; Lord et al., 2000) during diagnosis, with available scores. In 13 cases, the Autism Diagnostic Interview-Revised (ADI-R; Bölte & Poustka, 2001; Lord et al., 1994) had also been administered. Additionally, 7 autistic participants had a cooccurring diagnosis of attention deficit (hyperactivity) disorder (ADHD), 16 of depression, and 17 of anxiety.

Neurotypical (NT) participants were recruited from the Center for Brain and Cognition (Universitat Pompeu Fabra; UPF) database and were screened and excluded based on predefined criteria, including a history of neuro-psychological or neuro-psychiatric disorders, motor tics, epilepsy, ADHD, and seizures.

The groups were matched for chronological age, biological sex assigned at birth, self-identified gender, and non-verbal intelligence scores (measured with Raven’s Progressive Matrices 2; John & Raven, 2003; see Table 1). All participants provided prior written informed consent, and the protocol and data handling received approval of the Institutional Committee for Ethical Review of Projects at UPF (CIREP-UPF). Following the experiment, participants were debriefed and received 10Euro /hour in compensation for their time.

**Table 1.** Demographic and trait characteristics of subject samples in all groups. Count is provided for gender and sex, and means (with standard deviations) for all other items. F /M /O /I=female, male, other, intersexual, AQ=Autism Spectrum Quotient score, LSAS-SR=Liebowitz Social Anxiety Scale – Self Reported, R / A /L=Right, Ambidextrous, Left, OR=odds ratio in Fisher’s Exact Test. Statistically significant tests were marked with *** for p<.001. *Ns*=non-significant.

|  | AUT<br>N=32 | NT<br>N=34 | Group<br>comparisons<br>AUT vs. NT |
| --- | --- | --- | --- |
| **Gender** – F:M:O | 15:12:5 | 19:15:0 | ns |
| **Sex** – F:M:I | 20:12:0 | 19:15:0 | OR=1.31 |
| **Age** (years) | 32.2 (9.6) | 27.5 (10.8) | $t=-1.86$ |
| **AQ** (total) | 84.8 (10.7) | 57.6 (8.7) | $t=-11.28^{***}$ |
| **LSAS-SR** | 83.1 (25.9) | 40.8 (21.3) | $t=-7.23^{***}$ |
| **Handedness** – R:A:L | 31:1:0 | 33:0:1 | ns |
| **IQ** | 113.8 (13.3) | 111.4 (12.1) | $t=-0.78$ |
| **ADOS Total** (N=24) | 10.29 (5.22) | - | - |
| **ADOS Comm.** (N=23) | 3.35 (1.43) | - | - |
| **ADOS Social** (N=23) | 6.04 (2.4) | - | - |

### 2.3. Stimuli

The original stimuli for the preferential looking task, created by students of Marie Schaer (Geneva University), comprised 10 six-second-long videos featuring side-by-side silent videoclips, one of biological motion (social condition; SOC), depicting an actor producing meaningless facial gestures, and another of dynamic geometric motion (GEO). Side of SOC and GEO was equiprobable and randomly distributed across trials. We edited the original SOC stimuli (used in Polzer et al., 2022) replacing the original gradient grey background with solid black, to match the background in the GEO videos. This was done to limit the differences in average luminance, which may affect pupil sizes (e.g., McDougal & Gamlin, 2015). Luminance and contrast for each stimulus were nevertheless calculated (using custom Matlab code) and used as covariates in statistical analyses. For task 2, we created separate SOC and GEO videos featuring one single central stimulus, cropped out from the side-by-side videos.

### 2.4. Procedure

Two tasks were administered, during which eye gaze position and pupillary data were recorded via an eye-tracker (details below). Experiment duration was approximately 10 min. Participants were seated 50 cm from a 1920 × 1080 pixels HP Omen 25 computer monitor. In both tasks, participants were instructed to focus on the screen and watch a series of videos. Ten side-by-side SOC / GEO stimuli in task 1 (960 × 540 pixels, 28 × 16 degrees of visual angle) and 10 SOC and 10 GEO videos in task 2 (480 × 540 pixels, 14 × 16 degrees of visual angle) were presented in random order at the centre of a white screen. A two-second fixation cross preceded each stimulus presentation. Task order was fixed for all participants.

### 2.5. Apparatus and pre-processing

A Tobii Pro Spectrum eye tracker was utilised to binocularly record pupillary data and gaze behaviour at a rate of 120 Hz, without chinrest. Prior to the task, a standard 5-point calibration was performed. Offline pre-processing of eye data was conducted using custom R code following Kret & Sjak-Shie (2018). This involved blink and missing data interpolation, as well as filtering and smoothing using a moving average algorithm (30 ms window). Gaze position and pupil data were averaged across eyes. Data points where gaze was not focused on the stimuli were excluded. To enhance the computational efficiency of the analysis, we averaged the data over 50 ms bins, resulting in 20-Hz downsampling.

### 2.6. Data exclusion

Eye tracking data could not be collected from two participants due to calibration issues. Further, pupil datasets with over 50% rejected trials (i.e., containing more than 50% missing data due to blinks, gaze outside of stimuli, or poor data quality) were excluded from analyses (N=8). If a participant was rejected from at least one task, their datasets for both tasks were excluded.

### 2.7. Data analysis

Data analyses were performed using R 4.3.2 (R Core Team, 2023). Alpha-level for all tests was *p*=.05. Data and code are available at https://osf.io/nc7xm/ (including a rendered html file with all the analyses). Within the linear mixed model framework, all regression models were built with the *lme4* package 1.1.35.1 (Bates et al., 2015) and *lmerTest* package 3.1.3 (Kuznetsova et al., 2017). Post-hoc tests were adjusted for multiple comparisons with Holm-Bonferroni correction via the *glht* function (Hothorn et al., 2008). To approximate standard effect sizes, partial Cohen’s f (*f*_p_) and Cohen’s *d* were calculated with the *effectsize* package 0.8.6 (Ben-Shachar et al., 2020). Goodness of fit for mixed models was estimated with marginal and conditional *R*^*2*^ (Nakagawa & Schielzeth, 2013), in which marginal *R*^*2*^ (*R*^*2 m*^) reflects variance explained by fixed factors, and conditional *R*^*2*^ (*R*^*2 c*^) – variance explained by the entire model. The *p*-values were computed via Wald-statistics approximation. To assess the strength of evidence for null effects, Bayes Factors were calculated based on the Bayesian Information Criterion to evaluate the likelihood of the data occurring under a model without the hypothesised effect.

Two types of outcome variables were employed: looking preference (hypothesis 1), and pupil size (hypothesis 2). Looking preference was computed as the percentage of time gazing at SOC, relative to the total time (SOC plus GEO) in the trial. Scores greater than 0 indicate a preference for SOC (with 100 representing constant gaze on SOC), while scores lower than 0 suggest a preference for GEO (with -100 indicating constant gaze on GEO).

To examine hypothesis 1, a linear mixed model was constructed with looking preference as the dependent variable, group as predictor, and random intercepts for participants and trial. If group would significantly predict looking preference, so that AUT would be associated with lower looking preference scores than NT, we planned one-tailed t-tests per each group to assess significant deviations from 0.

For hypothesis 2, we examined the difference in pupil sizes between SOC and GEO (SOC – GEO) using growth curve analysis (Mirman, 2017). While our pre-registration specified starting the analysis from 1s after stimulus onset (excluding the initial light-evoked response), here we report the full window of stimulus presentation as this analysis allows modelling of the entire course of the pupil response. The overall time course of pupil responses was modelled with third-order (cubic) orthogonal polynomials. Compared to a null model (with only the intercept as a fixed predictor), models with random intercept for participants, and luminance and contrast as covariates, all demonstrated improved model fit and were therefore included in the model. If group was found to significantly influence the pupil response course, in the pre-registration we planned to build separate models for each group with condition as a predictor. However, to capture the time course of the pupillary response simultaneously across the two groups, we finally opted instead to 1) perform secondary pair-wise t-tests for the group effect in consecutive 500ms time bins of the pupil response, and 2) construct a secondary model with absolute (rather than relative) responses per condition.

## 3. RESULTS

### 3.1. Primary analyses

#### 3.1.1. Looking preference

Summary statistics and data distribution for looking preference across groups are shown in Table 2 and Figure 1 (top panel). As hypothesised (hypothesis 1), a model with random intercepts for participants and trials (model’s R^2m^ 0.03, R^2c^ 0.44) yielded a statistically significant effect of group on looking preference, *est*.=17.23, *SE*=8.18, *t*(65.9)=2.11, *p*=.04, *f*_p_=0.26. In the following post-hoc t-tests, we examined whether AUT show the hypothesised preference for GEO and NT for SOC (one-side tests against 0). These yielded a significant SOC preference in NT, *t*(33)=3.71, *p*_*corr*_ <.001, *d*=0.64, and no significant preference one way or another in AUT, *t*(31)=0.55, *p*_*corr*_ =.71, *d*=0.1.

**Table 2.** Preferential looking – summary statistics. Value of 0 signifies no looking preference for either motion type (half looking time in the trial spent gazing SOC and half GEO).

|  | Mean | SD | Min | Max | %<br>participants<br>showing<br>SOC<br>preference | %<br>participants<br>showing<br>GEO<br>preference |
| --- | --- | --- | --- | --- | --- | --- |
| AUT | 3.41 | 34.93 | -77.10 | 61.75 | 53% | 47% |
| NT | 20.64 | 32.43 | -60.82 | 88.14 | 82% | 18% |

**Figure 1.**
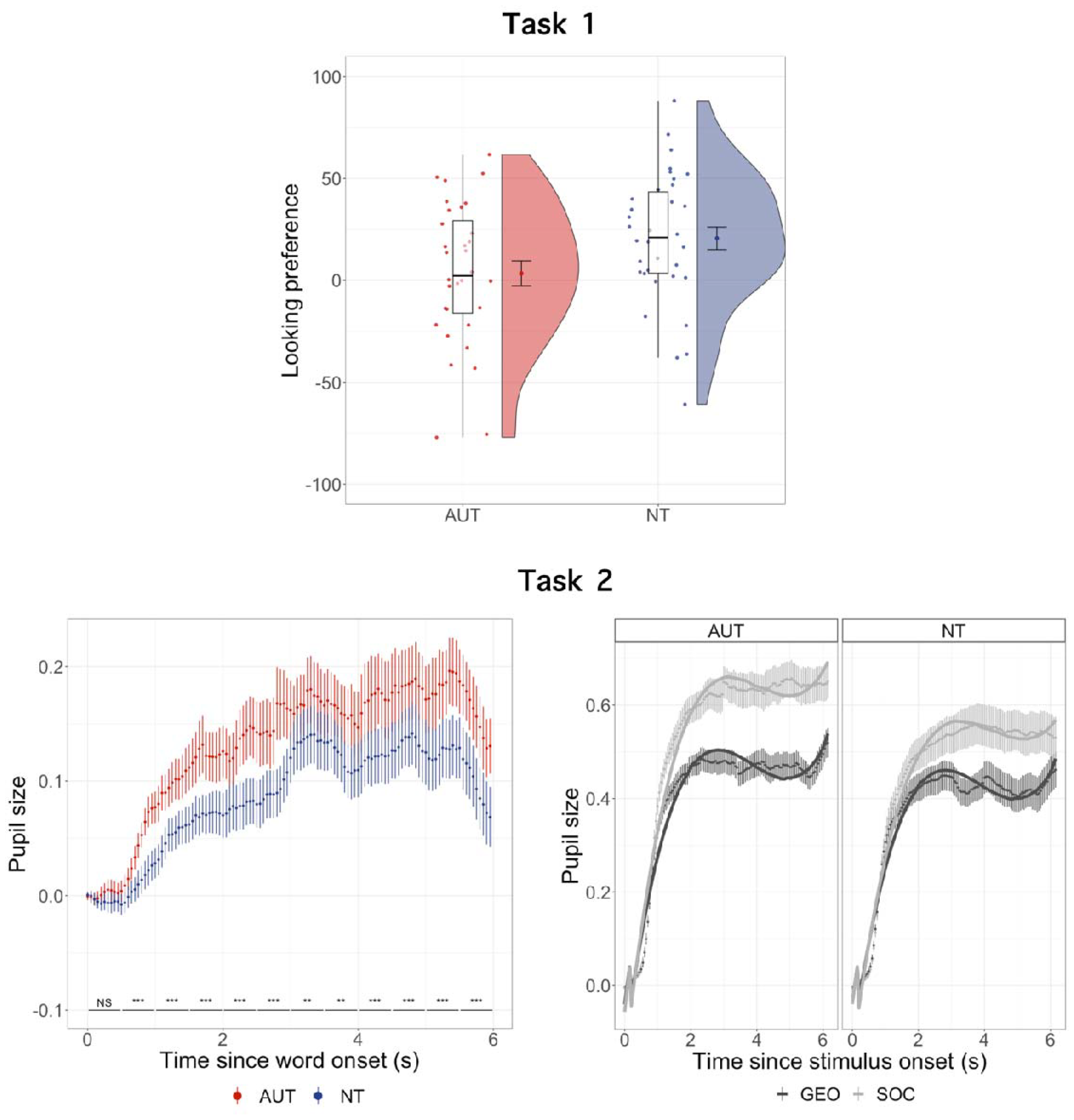
[Top panel] Looking preference across groups. Each point represents the average looking preference across trials for a participant. The more positive the value, the higher the preference for social motion (SOC). **[Bottom panel, left side] Relative pupil response to SOC vs. GEO across groups in the passive viewing task**. Line annotations in the bottom of the plot represent post-hoc t-tests for the effect of group in consecutive 500-ms bins (**=p<.01, ***=p<.001, NS=non-significant; all after Holm-Bonferroni correction). **[Bottom panel, right side] Pupil responses per condition and group in the passive viewing task**. Points illustrate observed data, with standard errors. Solid lines represent predicted responses by the secondary model with condition, group, and their interaction as predictors. Error bars denote standard errors in all plots.

#### 3.1.2. Pupillary responses

Pupillary responses in task 2 are shown in Figure 1 (bottom left panel). The primary pupil size model (model’s R^2^_m_=0.15, R^2^_c_=0.88) tested the effect of group on the relative response between conditions (SOC – GEO). The group had an effect on the cubic term, *est*=-0.79, *SE*=0.35, *t*(66.99)=-2.25, *p*=.03 (the effect size of the interaction of group and polynomials was *f*_p_=0.3). Together with visual inspection of Figure 1, the effect suggests faster and more strong pupil responses in AUT than in NT in response to SOC. There was also a trend effect of group on the intercept, *est*=-0.05, *SE*=0.03, *t*(66.02)=-1.72, *p*=.09, *f*_p_=0.21 (with larger pupil sizes for AUT than NT), which suggests that the groups do not differ in the overall pupil size.

To further interpret the group effects on the pupil responses, we performed secondary pair-wise t-tests for the group effect of the relative SOC-GEO pupil response in consecutive 500ms time bins (see Figure 1, annotations in the bottom left panel). In line with the primary model, except from the first bin (0 to 500 ms after stimulus onset), all the following bins showed significantly larger relative response to SOC vs. GEO in AUT than in NT (all *p*_*corr*_ • .005, *d*=0.13). Finally, to further visualise this group effect, we built a regression model with absolute (rather than relative) pupil responses to both motion types (SOC, GEO) across the groups (see Figure 1, bottom right panel), which yielded a significant interaction of group and condition, confirming larger pupil sizes in AUT than in NT in response to SOC (see section 1 in the Supplement).

### 3.2. Exploratory analyses

#### 3.2.1. Social anxiety traits in pupillary responses

Larger relative pupil responses to SOC in AUT than NT was an unexpected result. Hence, we further explored if social anxiety traits, measured with the Liebowitz Social Anxiety Scale (LSAS-SR; Liebowitz, 1987), could modulate this effect. However, social anxiety traits had no significant effect on the pupillary responses (all *p* • .25), their addition did not improve model’s fit, *X*^*2*^(8)=6.94, *p*=.54, and BF provided overwhelming evidence in favour of the model without them (BF01>1000).

#### 3.2.2. Pupil responses in the preferential looking task

We further investigated pupil responses in the preferential looking paradigm (task 1) by categorizing data points based on gaze location, distinguishing between SOC and GEO conditions. Employing the same analyses as described for task 2, data in task 1 revealed a comparable condition x group modulations of pupil responses (see section 2 in the Supplement).

#### 3.2.3. Covariates in the looking preference and pupil response models

We further explored whether age, biological sex, IQ, autistic traits (AQ scores; Baron-Cohen et al., 2001; Lugo-Marín et al., 2019), or ADOS score (in autistic participants only) predict looking preference and pupillary responses additionally to group (see section 3 in the Supplement). In the two tasks, the only significant effect was that of IQ interacting with group in the preferential looking task, so that higher IQ was linked to higher SOC preference in NT, but lower SOC preference in AUT. However, inclusion of IQ in the primary model was not supported by BF. No other additional predictor significantly improved the primary models of looking preference or pupil sizes. Additionally, we replaced the group with continuous AQ scores as a predictor in both primary models to explore whether quantified behavioural expressions of autism replicate the group effects. Autistic traits regardless of the diagnostic status predicted looking preference in task 1 (*p=*.04), but pupillary responses in task 2.

#### 3.2.4. Looking preference and pupil size correlations

We observed an interaction of pupil size and group on trial-by-trial looking preference. While in NT the pupil size did not predict looking preference, in AUT the larger the pupil size, the larger the social motion preference (see section 4 in the Supplement for details and a plot). We could not compute a looking preference in the passive viewing task.

#### 3.2.5. Predicting diagnosis status with the looking preference score

A logistic regression model did not provide evidence to support a significant association between looking preference score and the log-odds of the diagnostic status (see section 5 in the Supplement). Also, “social” and “geometric” autistic responders (those who showed relatively more time looking at SOC or GEO; Pierce et al., 2011) did not differ in any trait / clinical or demographic characteristics.

## 4. DISCUSSION

This study investigated whether autistic adults display characteristic looking preference and pupillary responses to social motion. We found that autistic individuals showed reduced spontaneous looking toward social stimuli, compared to neurotypicals. What is more, while the neurotypical group showed a strong imbalance for social motion, there was no clear preference for either motion type in the autistic group. This suggests that the atypical looking preferences present in autistic children persist into adulthood. The second, unexpected, finding was that pupillary responses in the autistic group showed faster and larger dilation for social motion than in the neurotypical group. We discuss these results in the light of the previous literature and our additional, exploratory analyses.

### 4.1. Reduced social motion looking preference in autism

Most neurotypical participants (82%) spent relatively more time looking at social than at geometric stimuli, leading to a strong group-level preference for social motion. Instead, the autistic group was evenly split, with 53% with social preference trend vs. 47% with geometric preference trend, and showed no statistically significant preference at the group level. This reduced interest in social motion in the autistic group is in line with previous literature using similar stimuli in children and adults (Del Valle Rubido et al., 2020; Franchini et al., 2016, 2017; Fujioka et al., 2016; Hong et al., 2019, 2019; Moore et al., 2018; Pierce et al., 2011, 2016; Polzer et al., 2022; Shaffer et al., 2017; Vargas-Cuentas et al., 2017; Wen et al., 2022). This pattern might be because of lack of social motivation and orienting in autism (Chevallier et al., 2012), because they find geometric motion more attractive, or both. Autistic individuals often show a bias towards observing repetitive patterns (Pierce et al., 2011) and display visuo-spatial strengths, like remembering geometric shapes (Hillier et al., 2007). Indeed, gaze patterns are more effective at discriminating between autistic and non-autistic children when contrasting faces versus geometric stimuli, as compared to assessing differences solely in the duration of gaze fixation on socially salient regions like the eyes (Kwon et al., 2019).

Aside from preferences, an alternative reason for the group differences in looking patterns might lie in the heterogeneity of the autistic sample in terms of (1) their responsiveness to the task or in terms of (2) autistic traits / symptom intensity. The first is not likely given that the standard deviation of the looking preference scores was similar between the groups (see Table 2) and the group difference seems to be accounted for by an average shift (see Figure 1, top panel). Indeed, looking preference scores in our study ranged between 11% and 81% for autism, and between 20% and 94% for neurotypical individuals, which aligns well with previous findings in children (Franchini et al., 2016, 2017; Pierce et al., 2011, 2016; Polzer et al., 2022).

The heterogenous looking preference scores may also vary with autistic traits /symptoms. Previous studies linked heterogeneity in the social motion preference to adaptive functioning in autistic children (Bacon et al., 2020; Franchini et al., 2016, 2017; Moore et al., 2018; Pierce et al., 2016). This supports the hypothesis that alterations in orienting and bias away from social motion may underlie social difficulties in autism. However, in our data neither symptom intensity (measured with ADOS) nor autistic traits (measured with AQ) predicted preferential looking or pupil responses in the autism group. At the same time, higher AQ scores across all subjects (autistic or not) were linked to smaller social motion preference when replacing group as a predictor. Together, this implies that social preference might not capture distinctive socio-communicative characteristics of autism in adulthood but rather reflects a general difference between autistic and non-autistic populations.

### 4.2. Looking preference as autism marker

Despite the significant group difference in relative looking preference, this measure failed to predict diagnosis status in an exploratory analysis. In their original article, Pierce et al. (2011) indicated that a cut-off of 69% of looking time on geometric motion yielded a 100% positive predictive value for autism diagnosis. This was further supported (98% specificity, 23-33% sensitivity) by a large-scale study from the same research group in over 1,400 toddlers (Wen et al., 2022). Just a visual inspection of Figure 1 reveals that our data do not show a similar potential.

One reason for the difference between our results and those of Pierce and colleagues is the nature of the stimuli. Our social stimuli consisted of actors producing a series of dynamically changing, non-prototypical, exaggerated facial expressions, which might be an unusual and less familiar motion than those used in previous studies, such as dancing people (e.g., Pierce et al., 2011) or playing children (e.g., Shaffer et al., 2017). Nevertheless, we observed a group difference, which suggests that the lower social motion preference in autism is a robust effect across various stimuli characteristics, albeit less pronounced in our stimuli.

Another reason for looking preference not predicting diagnosis in our study may be the age of the participants. Few studies, including this one, have investigated whether a preference for complex social motion can serve as a marker of autism in adults. More data are needed before disregarding this as a potential marker, especially in the light of some promising results in children (Moore et al., 2018; Wen et al., 2022). Because children and adults differ in gaze behaviour in free looking tasks (Franchak et al., 2016), it is possible that social vs. geometric motion looking preference is more indicative of autism in younger samples. In this study, age (19-56 years) did not significantly impact preferential looking, consistent with a meta-analysis of biomotion processing (Federici et al., 2020), and a well-powered study across a younger but broad age range (6-30 years; Mason et al., 2021). However, a meta-analysis of biomotion studies in autism by Todorova et al. (2019) found larger effect sizes for the group difference in younger ages, suggesting compensatory strategies in adults. It is thus possible that autistic and neurotypical individuals become more similar in viewing behaviour over time. Together, given the large variance in preferential looking scores of the autistic group in our study, we view the observed lower social motion preference in autism as indicative of a difference rather than a deficit in social attention (Mason et al., 2021).

### 4.3. Pupillary responses to social vs. geometric motion in autism

To explore mechanisms underlying lowered social motion preference in autistic compared to neurotypical adults, we investigated pupillary responses. Prior research in children has associated salience responses within the LC-NE system, as measured by pupillary responses, with differences in social attention observed in autism (Polzer et al., 2022). In our study, while both groups showed larger pupil responses to social than geometric stimuli, these responses were faster and larger in the autism group. This unexpected finding did not support the predicted pattern (larger pupil sizes to geometric motion in the autistic adults and to social motion in the neurotypical adults), based on previously reported results in pre-schoolers (Polzer et al., 2022). Given that social anxiety is commonly cooccurring in autism (Spain et al., 2018), one potential interpretation was that the increased pupil responses to social motion in the autism group reflected heightened arousal towards social stimuli that could not be averted, at least by instruction. However, a secondary analysis revealed that the group effects could not be attributed to social anxiety traits.

Pupil size can also be a measure of subjective task difficulty and mental effort (Eckstein et al., 2017), as the LC-NE system controls cognitive functioning (Ramos & Arnsten, 2007). In this view, one could speculate that social motion required more cognitive effort than the geometric motion for both groups, but especially for the autistic participants. Our participants were instructed to not look away from the stimuli or avert their gaze, and so it might have been difficult to downregulate processing of the stimulus. This interpretation is consistent with that of Del Valle Rubido et al. (2020), who reported larger tonic pupil dilation in autistic compared to non-autistic adults in a social vs. geometric motion task. They interpreted these findings as indicative of heightened mental effort among autistic adults during social scene interpretation. Although they limited the analysis to tonic pupil changes, a similar pattern characterises the baseline-corrected phasic pupil responses in our study.

The finding of larger pupil responses to social motion in autistic adults stands in contrast with Polzer et al.’s (2022) results in children. Polzer and colleagues reported smaller phasic pupil dilation to social motion and larger dilation to geometric motion in autistic children, when watching them side-by-side. They interpreted this finding either as an alteration in bottom-up processing of sensory information, or as a differential higher-order attribution of salience. In this view, geometric motion, being more visually appealing, was perceived as having greater salience than social motion and /or its sensory processing was facilitated in autistic children. While our primary pupil response measurements where with centred images (task 2), we also found a similar pattern of results (with larger pupil sizes for social stimuli in autism; see section 2 in Supplement) when measuring pupil size in side-by-side presentations during the preferential looking task, thus comparable to Polzer et al.’s (2022). Following their account, our results would have to suggest heightened salience attributed to social stimuli in autistic individuals. Speculatively, autistic adults may employ compensatory strategies in social interpretation compared to children, resulting from learned experiences throughout life, with the brain adapting by assigning greater importance to social information to enhance processing efficiency.

With few studies reporting pupil responses to social vs. geometric motion in autism, definite interpretation of the underlying cognitive mechanisms would be premature. Nevertheless, it should be noted that our study introduces a significant methodological distinction when compared to previous studies (Del Valle Rubido et al., 2020; Polzer et al., 2022). By analysing pupil responses to centrally presented single stimuli, rather than the side-by-side, we addressed the issue of unequal data points per group and condition, and potential voluntary control of input.

### 4.4. Conclusions

In this study, we investigated social vs. geometric motion preference in autistic and neurotypical adults. While the neurotypical group exhibited a clear preference for social motion, the autistic group displayed no distinct preference but demonstrated faster and stronger pupil dilation to social motion. These findings contribute to the literature on reduced preference for social motion in autism and offer insights into the underlying cognitive processes, indicating potential increased cognitive effort and /or compensatory strategies when interpreting social motion in autism.

## Data Availability

The data and analysis code will be made publicly available upon publication at https://osf.io/nc7xm/.

## Acknowledgements

The authors are grateful to the participants who volunteered to take part in the study and offered their feedback which continues to help us improve this and future study designs, and to Marie Schaer who gave her permission to use the stimuli created in her laboratory.

