## Supplementary_material for "Preference for Social Motion in Autistic Adults"

Supplement
to
**Social motion preference in autism**
by Matyjek, Bast, and Soto Faraco

### 1. Secondary model of SOC vs. GEO in AUT and in NT in Task 2.

To further visualise the group effect found in the primary model testing hypothesis 2, we built a regression model with absolute (rather than relative) pupil responses to both motion types (SOC, GEO) across the groups. This model ($R^2_m$ = 0.49, $R^2_c$ = 0.91) revealed a main effect of condition, *est* = 0.15, *SE* = 0.002, *t*(16103.93) = 78.15, $p < .001$, $f_p = 0.69$, but not group, *est* = -0.04, *SE* = 0.04, *t*(66.52) = -1.03, *p* = .31, $f_p = 0.2$. Most importantly, there was an interaction effect of group and condition, *est* = -0.04, *SE* = 0.002, *t*(16103.93) = -18.59, $p < .001$, $f_p = 0.15$, and three-way interactions of group, condition, and all three time polynomials; linear: *est* = -1.5, *SE* = 0.3, *t*(16103.93) = -4.91, $p < .001$; quadratic: *est* = 0.08, *SE* = 0.3, *t*(16103.93) = 2.76, *p* = .006; and cubic: *est* = -1.13, *SE* = 0.3, *t*(16103.93) = -3.73, $p < .001$ (overall effect size for the three-way interaction: 0.05). These effects are in line with the group effect on the cubic term in the primary model and suggest that the groups differed in their pupil responses to SOC and GEO, so that the pupil dilated faster and more in AUT than in NT in response to SOC. Additionally, the group did not have a main effect on the polynomials (all $p \geq 0.35$), which suggests that the general shape of pupil response is similar between the groups. However, the condition had a significant effect on all the polynomials, showing that the responses to SOC are more pronounced than to GEO (all $p < .02$).

### 2. Pupillary responses in Task 1.

We examined pupil responses in Task 1 using the same analysis applied to Task 2 data. The results are presented here, with a cautionary note on the interpretation of statistical significance, given the exploratory nature of this analysis. Aggregated pupillary responses in Task 1 are shown in Supp. Fig. 1 (bottom left panel). The model (model's $R^2_m$ = 0.04, $R^2_c$ = 0.42) revealed that the linear term was significant and positive, describing larger pupil sizes with time, *est* = 2.18, *SE* = 0.77, *t*(66.57) = 2.85, *p* = .006, and a cubic effect starting with an increase. The quadratic term and cubit terms were negative and not significant (both *ps* >= 0.24). Most importantly, the group had an effect on the linear term, *est* = -2.54, *SE* = 1.06, *t*(66.04) = -2.39, *p* = .02, effect size of the interaction of group and polynomials was $f_p$ = 0.3. However, as one would expect, the data exhibited more noise, and some effects observed in the second task did not achieve significance (although significance should be interpreted cautiously in exploratory analyses). Notably, while AUT, in comparison to NT, showed a trend of larger pupil responses to SOC than GEO, this effect emerged around four seconds after stimulus onset, contrasting with the 0.5-second onset observed in the passive looking task.

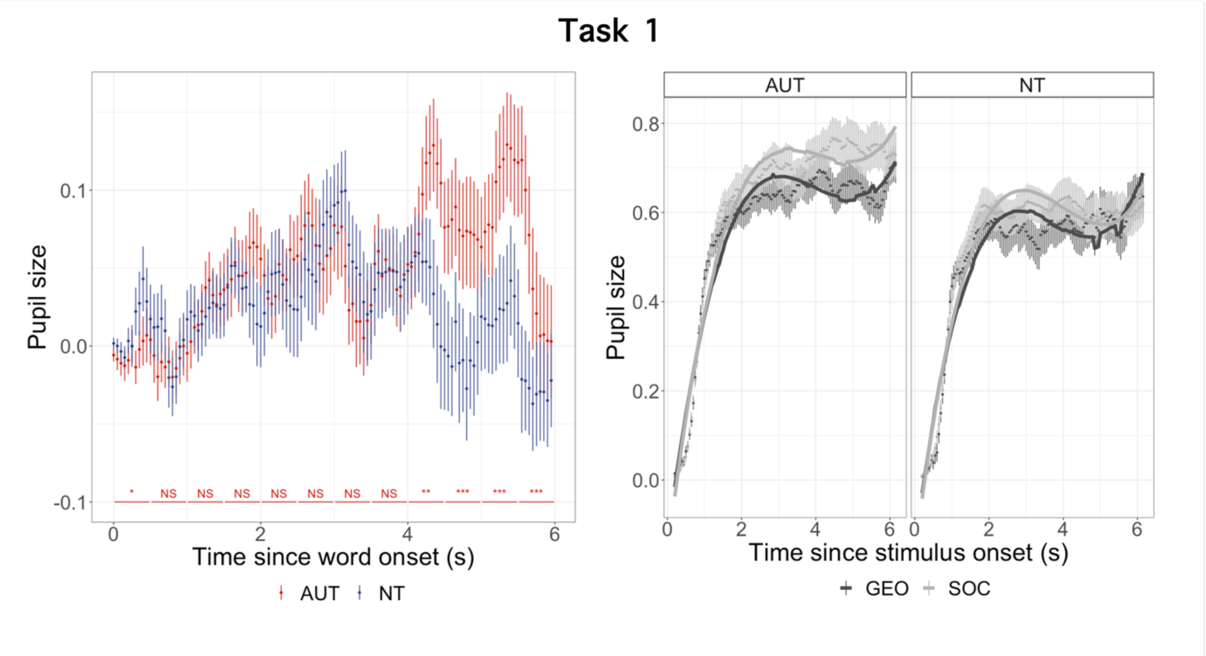


Supp. Fig. 1 **[Bottom panel, left side] Relative pupil response to SOC vs. GEO across groups in Task 1.** Error bars denote standard errors. Annotations in the bottom of the plot represent post-hoc significance of t-tests for the effect of group in consecutive 500-ms bins (* = p < .05, ** = p < .01, *** = p < .001, NS = non-significant; all after Holm-Bonferroni correction). **[Bottom panel, right side] Pupil responses per condition and group in Task 1.** Points illustrate observed data, with standard errors. Solid lines represent predicted responses by the secondary model with condition, group, and their interaction as predictors.

### 3. Exploratory covariates in looking preference and pupil response models

In the preferential looking data (Task 1), IQ significantly interacted with group, *est* = 18.71, *SE* = 7.91, $t(65.88) = 2.36$, $p = .02$, $f_p = 0.29$, so that higher IQ was linked to higher SOC preference in NT, but lower SOC preference in AUT (see Supp. Fig. 2). However, BF provided strong evidence against including IQ in the primary model (BF01 = 44). No other additional predictor had a significant effect on looking preference (all $p \geq 0.08$) or improved the model's fit, all $X^2(2) \leq 5.39$, all $p \geq .19$, all BF01 ≥ 124. In pupillary data (Task 2) no predictor was significant (all p ≥ 0.09), or improved the model's fit, all $X^2(8) \leq 8.04$, all $p \geq .43$, all BF01 > 1000.

When replacing the group for the continuous AQ score as a predictor in the primary models, AQ significantly predicted looking preference in Task 1: *est* = -0.54, *SE* = 0.25, $t(65.89) = -2.18$, $p = .04$, $f_p = 0.27$, but not the pupil sizes in either term in Task 2 ($p >= .35$).

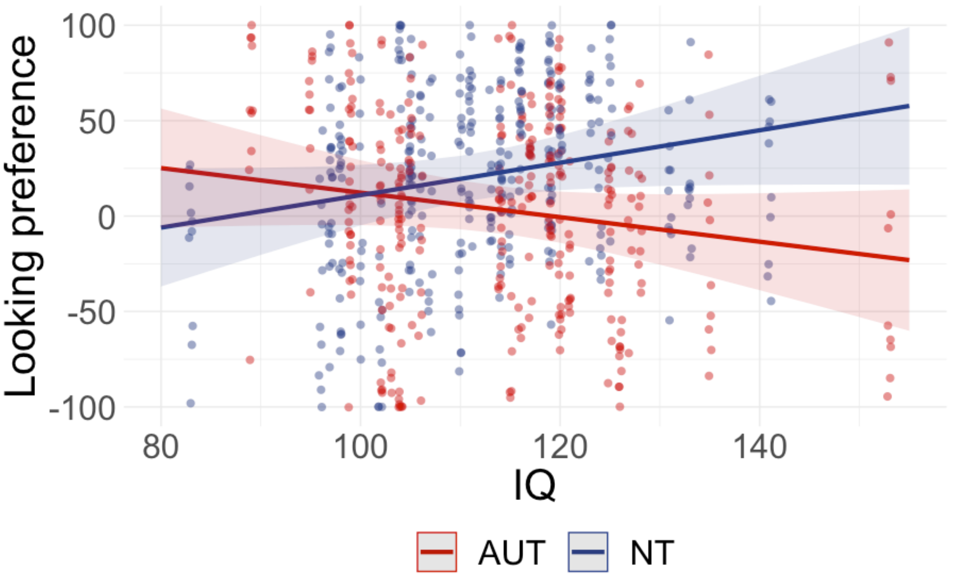


Supp. Fig. 2 **Interaction of group and IQ in looking preference** (Task 1).

### 4. Looking preference – pupil size correlation

In Task 1, the looking preference score in each trial was significantly predicted by the pupil size, *est* = 70.68, *SE* = 16.24, *t*(619.83) = 4.35, $p < .001$, group, *est* = 48.15, *SE* = 15.17, *t*(250.88) = 3.17, *p* = .002, and interaction of group and pupil size, *est* = -51.80, *SE* = 22.51, *t*(598.09) = -2.3, *p* = .02. The plot of these effects in Supp. Fig. 3 suggest that while in NT the pupil size does not predict looking preference, in AUT the larger the pupil size, the larger the social motion preference.

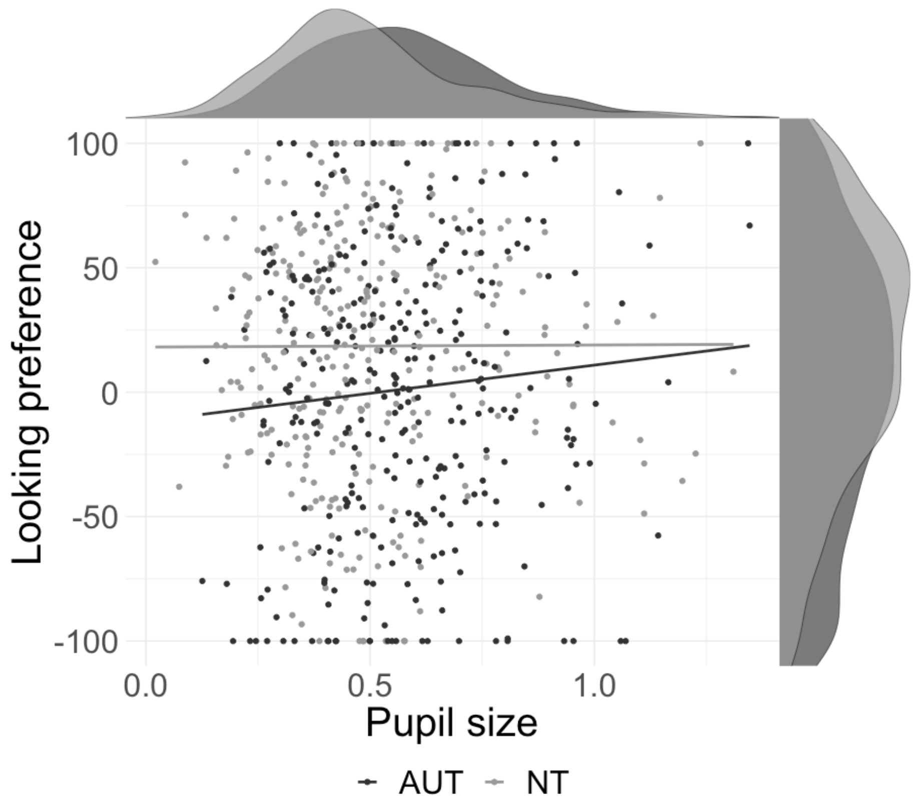


Supp. Fig. 3 **Correlations of the looking preference and pupil size in Task 1.**

### 5. Predicting diagnosis status with the looking preference score

To compare the accuracy of responses between conditions, we built a generalised linear mixed model (GLMM) with a binary dependent variable for group (or: participant's diagnostic status; 1 = AUT, 0 = NT) and looking preference score as a single predictor. Random intercepts for participants were included. The logistic regression analysis revealed that the intercept was 13.61 ($z = 6.932$, $p < .001$), indicating the log-odds of the outcome when looking preference is zero. The coefficient for the scaled looking preference variable was 0.24 ($SE = 0.91$, $z = 0.265$, $p = 0.791$), suggesting a non-significant effect.

Then, following the analysis in the original paper by Pierce et al. (2011), we explored if the "social" and "geometric" autistic responders (i.e., those who showed relatively more time looking at SOC or GEO) differed in any trait/clinical or demographic characteristics. For that we conducted independent two-tailed t-tests between "social" and "geometric" autistic responders for age, IQ, AQ, ADOS total score, ADOS communication score, and ADOS social interaction score. No effect remained significant after an FDR correction, all *p*s > .98.
